# The Institutional and Cultural Context of Cross-National Variation in COVID-19 Outbreaks

**DOI:** 10.1101/2020.03.30.20047589

**Authors:** Wolfgang Messner

## Abstract

**Background:** The COVID-19 pandemic poses an unprecedented and cascading threat to the health and economic prosperity of the world’s population.

**Objectives:** To understand whether the institutional and cultural context influences the COVID-19 outbreak.

**Methods:** At the ecological level, regression coefficients are examined to figure out contextual variables influencing the pandemic’s exponential growth rate across 96 countries.

**Results:** While a strong institutional context is negatively associated with the outbreak (B = −0.55 … −0.64, *p* < 0.001), the pandemic’s growth rate is steeper in countries with a quality education system (B = 0.33, *p* < 0.001). Countries with an older population are more affected (B = 0.46, *p* < 0.001). Societies with individualistic (rather than collectivistic) values experience a flatter rate of pathogen proliferation (B = −0.31, *p* < 0.001), similarly for higher levels of power distance (B = −0.32, *p* < 0.001). Hedonistic values, that is seeking indulgence and not enduring restraints, are positively related to the outbreak (B = 0.23, p = 0.001).

**Conclusions:** The results emphasize the need for public policy makers to pay close attention to the institutional and cultural context in their respective countries when instigating measures aimed at constricting the pandemic’s growth.

## Introduction

As of March 21, 2020, more than 271364 cases of coronavirus disease 2019 (COVID-19) were confirmed worldwide. Italy, then the second most impacted country with 47021 confirmed cases, recorded its first three cases only on January 31, 2020.^1^ Efforts to completely contain the new virus largely failed. As a consequence of global mobility and trade, people carrying the virus arrive in countries without ongoing transmission. Governments are currently scrambling to put in unprecedented measures to flatten the curve, because the faster the infection curve rises, the quicker the national health care systems get overloaded beyond their capacity of treating people effectively. While ultimately the same number of people are likely to get infected, reducing the initial number of cases would make the outbreak easier to control overall.^2^

In this study, I examine cross-national variation in COVID-19 outbreaks in 96 countries to analyze the impact of global connectivity, national institutions, socio-demographic characteristics, and cultural values on the initial arc of the curve. While getting to know the epidemic through, inter alia, mathematical models is important for national and international countermeasures, experience from HIV shows that politics and ideology are often far more influential than evidence and best practice guidance.^3^ It is well acknowledged that politics is central to policy-making in health generally, and that the institutional and cultural context plays a defining role in health policy outcomes. With the H1N1 2009 influenza pandemic, social determinants of health affected outcomes beyond clinically recognized risk factors.^4^ Thus, getting to know the national context of the COVID-19 pandemic will be essential in informing the development of evidence-based measures.

## Model and method

I implemented a linear regression model, in which the exponential growth rate of confirmed COVID-19 cases is regressed on institutional, socio-demographic, and cultural variables associated with testing and reporting cases, supporting the pathogen’s path, and controlling the outbreak. As of March 21, 2020, there is sufficient COVID-19 outbreak data to estimate the model for 96 countries. All variables are detailed in Table 1. The aim of this ecological approach is to study health in an environmental context.^5^

**Table 1:**
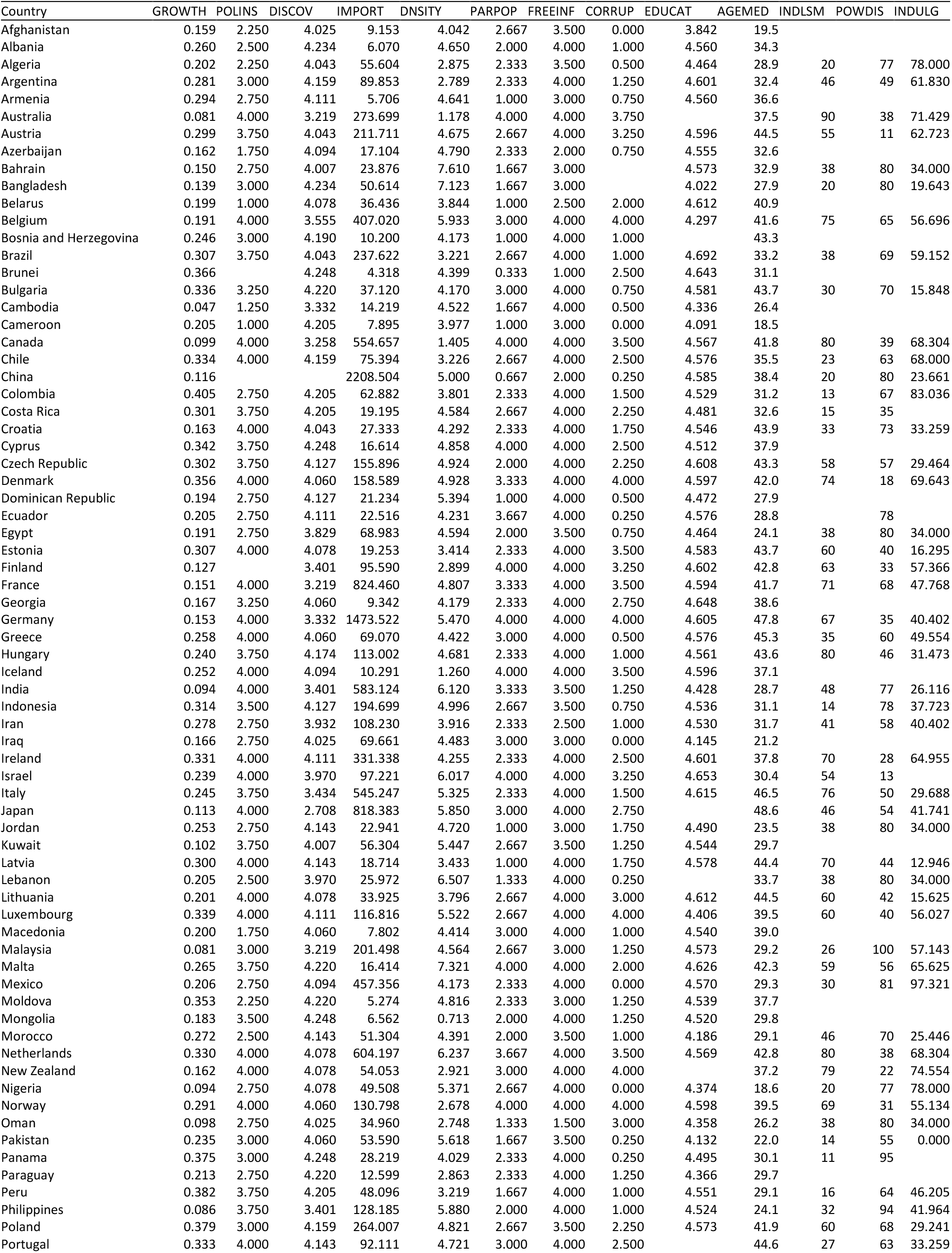
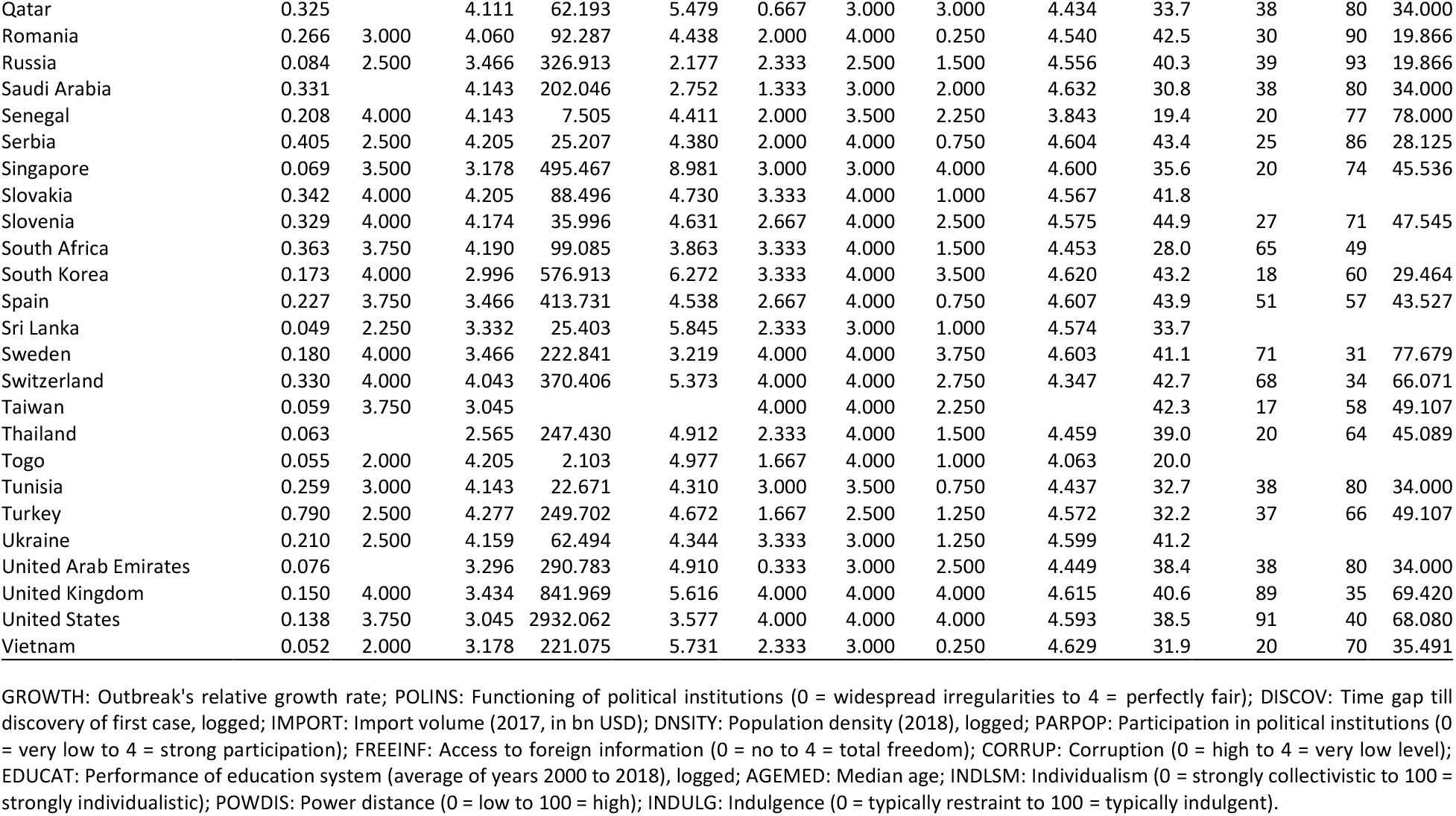
Relative growth rate of COVID-19 and contextual variables

### Outbreak data

In this study, I use data from the European Center for Disease Control and Prevention (ECDC), which is an EU agency established in 2005 with the aim to strengthen Europe’s defense against infectious diseases. The ECDC collects and harmonizes data from around the world, thus providing a global perspective on the evolving pandemic; the datafile is available via Our World in Data, an effort by the University of Oxford and Global Change Data Lab.^1^ Note that the World Health Organization (WHO) changed their cutoff time on March 18, 2020, and, due to overlaps, their data is not suitable for understanding the pandemic’s development over time beyond this date.^1^ To have enough datapoints for estimating the relative growth rate (dependent variable GROWTH) in an exponential population model, I only include countries which have reported their first case on or before March 12, 2020, as per the ECDC dataset. With a change point analysis using the Fisher discriminant ratio as a kernel function, I confirm that the first reporting date is in fact the start of the outbreak.^6^ Accordingly, there are no later significant change points in the outbreak.

### Testing and reporting cases

During the current COVID-19 outbreak, practically all countries are struggling to test every person who should be tested from a medical standpoint. Under the guidelines of most countries, clinicians will test suspected patients only if they have travelled to an epidemic region.^7^ The more tests a country performs, the more confirmed cases it tends to have. Because data on the number of tests performed is neither comparable across countries (it may refer to tests or individuals) nor updated regularly,^1^ I introduce variables into the regression model, which could purportedly be associated with a country’s capability and commitment to test and report. First, I use a perception indicator about the functioning of political institutions (independent variable POLINS; 0 = widespread irregularities to 4 = perfectly fair) from the 2016 edition of the International Profiles Database (IPD), which is a survey conducted by the French Directorate General of the Treasury.^8^ Second, I calculate the time between Jan 01, 2020 (as a rather random starting point) and discovering the first case (independent variable DISCOV). This time lag helps a country to learn from others’ experiences, and ramp up their own testing capabilities. As this is likely a non-linear effect, I logarithmically transform this measure in the regression model.

### Interconnectivity between populations

Because international connectivity between countries increases the potential spread of a pathogen,^9^ I introduce the independent variable IMPORT, which represents the value of all goods and other market services received by a country from the rest of the world (year 2017; in bn USD; based on data from the World Bank).^10^ Additionally, with the logged variable DNSITY, I capture a country’s population density, which is defined as all residents in a country divided by land area in square kilometers (year 2018; data from the World Bank).^11^

### Institutional context

Because strong stakeholder processes can bring benefits to accepting decisions being made by the government,^12,13^ I use an indicator on participation of the population in political institutions from the IPD database (independent variable PARPOP; 0 = very low to 4 = strong participation).^8^ Second, the society’s openness can be described by the freedom of access to foreign information (independent variable FREEINF; 0 = no to 4 = total freedom; from IPD).^8^ Third, the functioning of the public administration is, inter alia, mirrored in the level of corruption (independent variable CORRUP; 0 = high to 4 = very low level of corruption; from IPD).^8,14^

### Socio-demographic mapping

Variable EDUCAT is a logged indicator of an education system’s performance, calculated as the gross intake ratio to the last grade of primary education (average of years 2000 to 2018; data from the World Bank).^15^ And as older people (especially in Italy) seem to get hit more frequently by COVID-19,^16^ I introduce AGEMED as an independent variable for a country’s median age (current data from the CIA World Factbook).^17^

### Cultural variables

Given that culture determines the values and behaviors of societal members,^18^ specific behavioral manifestations of culture can influence the transmission of pathogens.^19^ Although country boundaries are not strictly synonymous with cultural boundaries, there is abundant evidence that geopolitical regions can serve as useful proxies for culture.^19^ Thus, I use scores from Hofstede’s dimensional framework of culture,^18^ available for 73 countries included in my analysis. Individualism (independent variable INDLSM, score of 1 to 100) is defined as a preference for a loosely-knit social framework, whereas collectivism (low scores on the same variable) represents a preference for a tightly-knit framework, in which individuals expect members of a particular ingroup to look after each other in exchange for unquestioning loyalty. Previous studies have shown that the regional prevalence of pathogens is negatively associated with individualism.^19^ Power distance (independent variable POWDIS, score of 1 to 100) expresses the degree to which the less powerful members of a society accept and expect that power is distributed unequally, with the fundamental issue being how societies handle inequalities among its members. Accordingly, the norm in countries with high values of POWDIS is the belief that everyone should have a defined place within the social order. The epidemiology of infections has been shown to be linked to power distance, but results are not conclusive.^20^ In low power distance cultures, people are less willing to accept directions from superiors,^21^ with potentially detrimental effects on controlling the outbreak of a pandemic. Conversely, in consumer research, country-level high power distance results in weaker perceptions of responsibility to aid others in a charitable way.^22^ Lastly, the dimension of indulgence (independent variable INDULG, score of 1 to 100) reflects hedonistic societies that allow people to enjoy life and have fun, as compared to societies where restraint is emphasized. It can be assumed that countries scoring high on the indulgence dimension will have more difficulty constraining social activity, implementing social distancing measures, and thereby restricting its citizens’ satisfying activities.

### Statistical results

To test the association of the context variables on the growth rate of COVID-19, I use linear regression with pairwise exclusion of missing values. The results suggest that a significant proportion of the total variation of the outbreak can be explained by the context variables, *F*(12,55) = 26.16, *p* < 0.001. Multiple *R*^*2*^ indicates that 85.09% of the variation in growth can be predicted by the context variables; estimated power to predict multiple *R*^*2*^ is at the maximum of 1.000, as calculated with G*Power 3.1. Table 2 expounds the regression coefficients. Multicollinearity in epidemiological studies can be a serious problem, being a result of unrepresentative samples or insufficient information in samples, that is not enough countries or omission of relevant variables.^23^ I have conducted several diagnostics to eliminate multicollinearity issues in the regression analysis. First, the VIF never exceeds 4 (see Table 2), which is well below the recommended threshold of 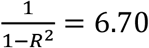. Second, the highest correlation coefficient is 0.683 between variables DISCOV and IMPORT, which is below the typical cutoff of 0.8. Only another two correlation coefficients are above the 0.5 cutoff (EDUCAT and AGEMED: −0.56; POLINS and FEEINF: −0.58). Third, the variance-decomposition matrix does not show any groups of predictors with high values. In summary, a multicollinearity problem can be excluded.

**Table 2:**
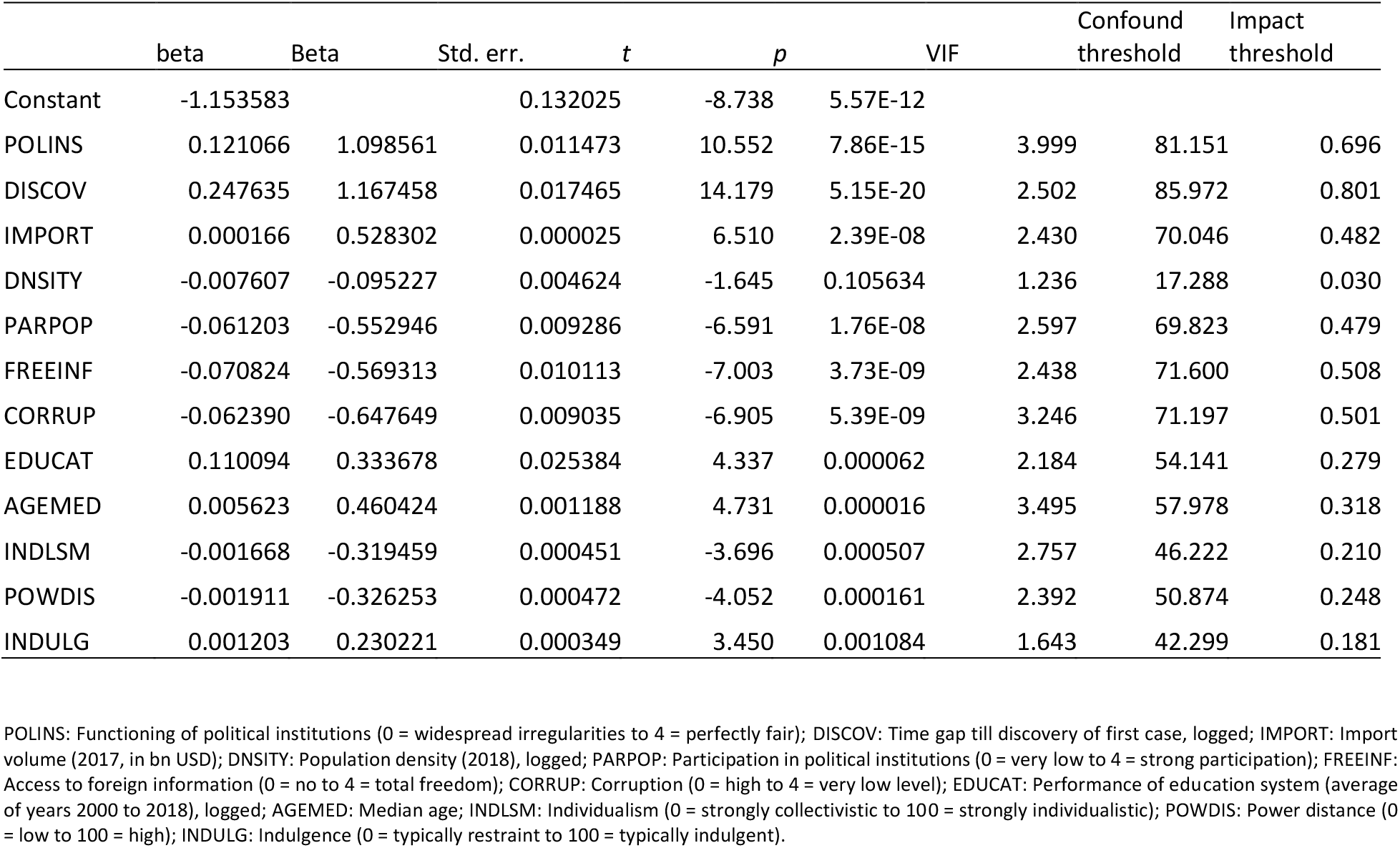
Regression results

Further, I conduct several tests to assess the robustness of the results by including other contextual variables. But because it is nearly impossible to establish a complete list of such confounding variables, I additionally quantify the potential impact of unobserved confounds (Table 2, column Impact threshold).^24^ For instance, the necessary impact of such a confound for the variable DISCOV would be 0.80, that is, to invalidate the inference that the time lag has on the growth rate, a confounding variable would have to be correlated with both GROWTH and DISCOV at 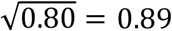, which is a strong correlation. Next, to alleviate concerns that the worldwide spread of the virus is not yet fully known and that this study might have been conducted too early in the pandemic, I ask how many countries would have to be replaced with unobserved cases for which the null hypothesis is true (i.e., the contextual variables have no influence on the growth rate) in order to invalidate the inference.^25^ As Table 2 (column Confound threshold) shows, about 86% of the countries would have to be replaced with countries for which the effect is zero in order to invalidate the influence of DISCOV. In summary, it can be claimed that the influence of the identified contextual variables on the pandemic’s growth rate is reasonably robust.

## Discussion

As expected, countries with functioning political institutions (POLINS) report a higher relative growth rate of the outbreak, probably due to a better testing and reporting infrastructure. Likewise, for countries that have been hit by the outbreak at a later point of time (DISCOV). The scatterplot in Figure 1 graphically depicts the relationship between discovery of the first case and the rate of the outbreak. In this diagram every dot represents a country; Turkey shows up as an outlier having reported their first case only on March 12, 2020,^2^ but showing a very rapid outbreak. International connectivity as measured by a country’s import volume (IMPORT) elevates the growth rate (Figure 2). Contrary to expectations, population density (DNSITY) is negatively related to the outbreak. Maybe people in densely populated countries are more likely to adhere to precautionary measures because they realize the danger of physical closeness to pathogen transmission?^26^ Or does this indicate that social distancing measures are more effective in crowded places? Yet, the DNSITY coefficient is not statistically significant in the regression model, and the confound threshold is rather low (*p* = 0.105, confound threshold 17.28%). A strong institutional context is negatively associated with the outbreak, as measured by participation in political institutions (PARPOP), access to foreign information (FREEINF), and absence of corruption (CORRUP). Rather surprisingly and contrary to the experience with HIV,^27^ the quality of a country’s education system is positively associated with the outbreak. Do people believe that the pathogen affects only poor countries, and therefore do not take precautionary measures seriously? Or do better educated people test more due to increased awareness? Providing a conclusive reasoning at this point in the COVID-19 outbreak is not possible, and I encourage further research in the months or years to come.

**Figure 1:**
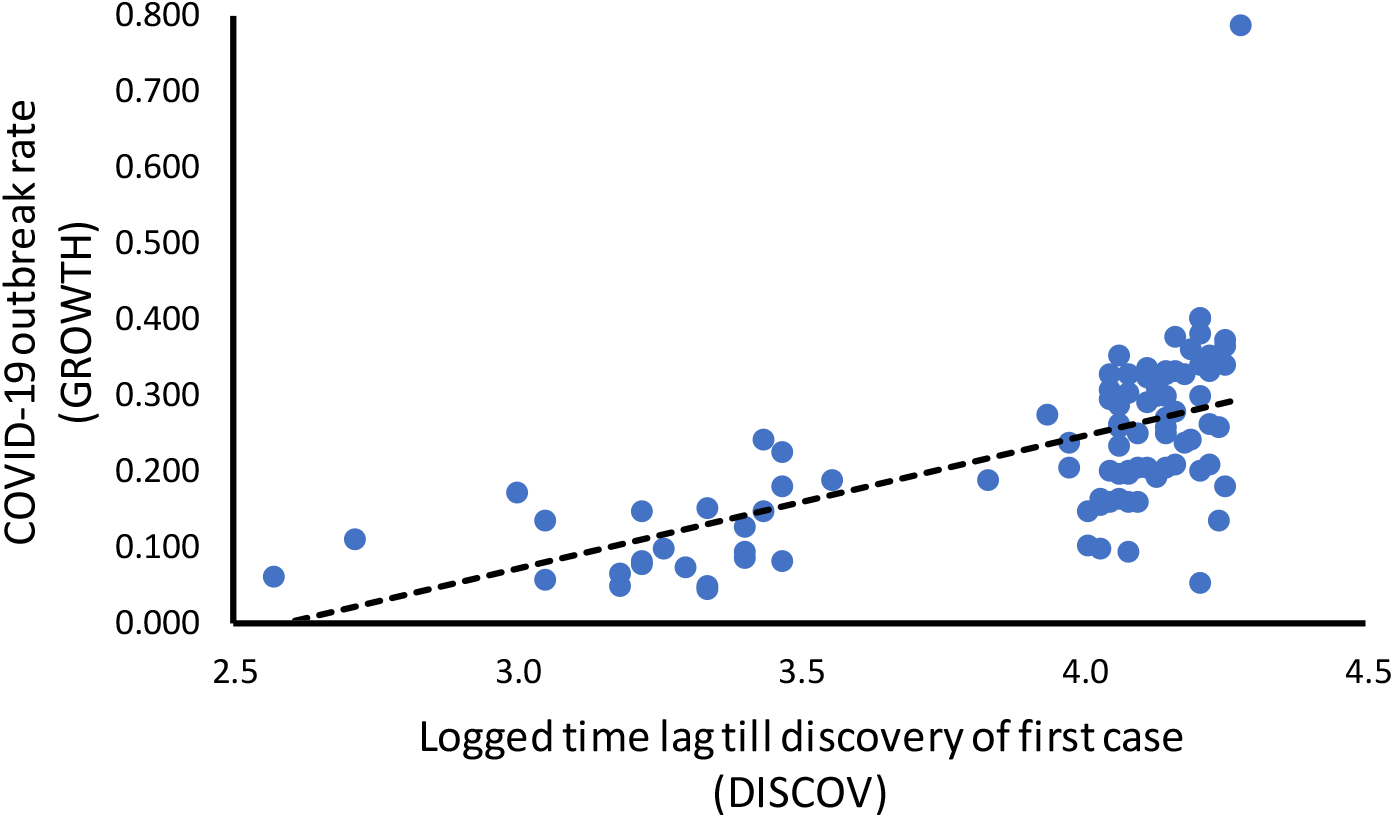
Association between time lag of COVID-19 outbreak and growth rate.

**Figure 2:**
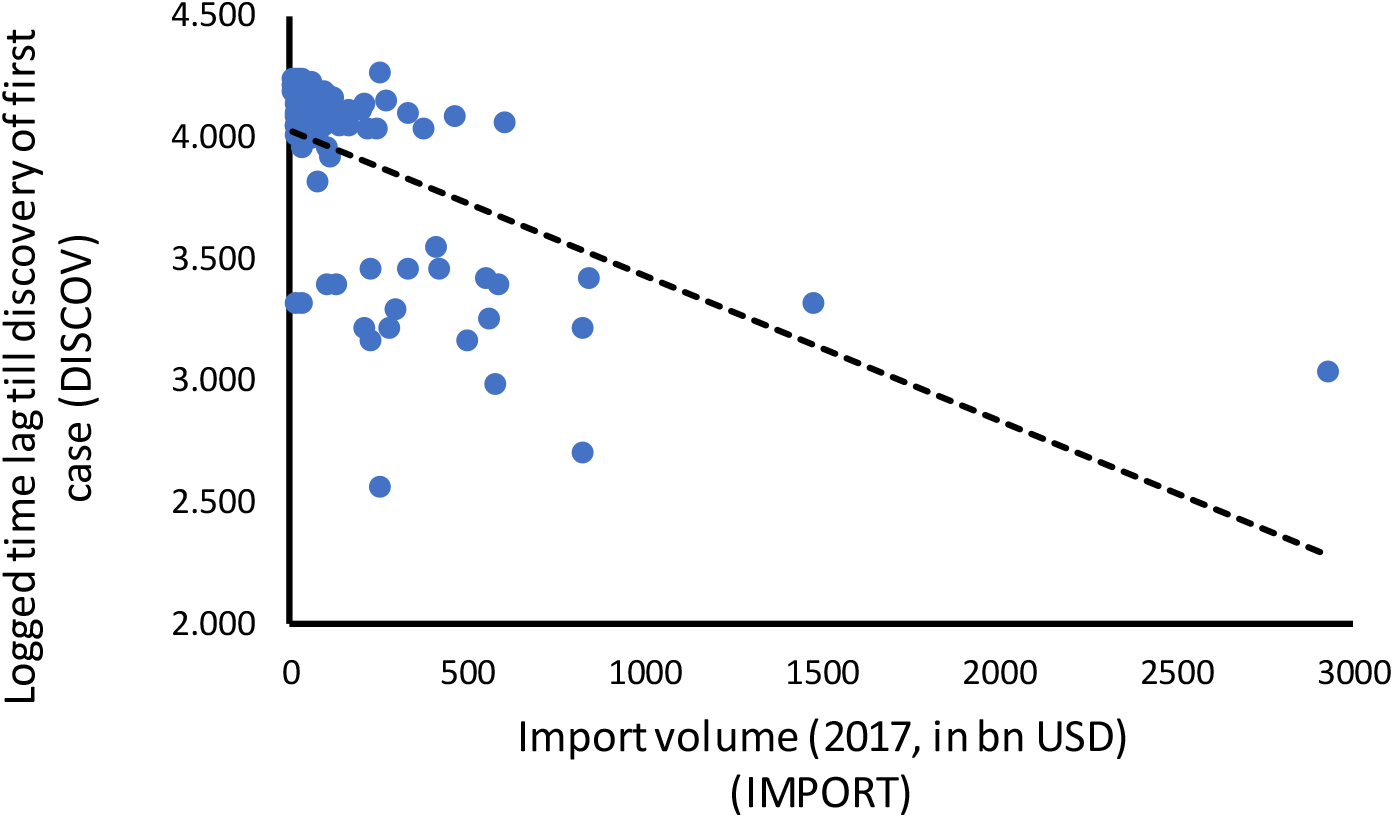
Association between import volume and time lag of COVID-19 outbreak.

Whilst potentially controversial, an association between cultural characteristics and the outbreak of the pandemic should not be totally surprising, since implementing countermeasures is ultimately behavioral science.^28^ The data shows that individualistic societies experience a lower outbreak growth rate, which is in line with previous studies about pathogen proliferation.^19^ People in more collectivistic cultures apparently find it more difficult to engage in social distancing practices. And because the effectiveness of social distancing measures has rarely been assessed before,^26^ this calls for a cross-cultural investigation in further research. Higher levels of power distance are associated with a lesser growth rate of the outbreak; it appears that individuals in low power distance cultures are less willing to blindly accept directions from the government on how to change their social behavior.^21^ Instead, they prefer a say in decisions affecting their lifestyle. Even though managing individuals’ obstinate behavior is quite a challenge in a pandemic, politicians in low-power distance countries need to work more towards achieving a buy-in of their electorate. Lastly, a country’s hedonistic tendency towards indulgence and not accepting restraints is positively linked to the outbreak.

My study indicates that governments need to tailor their strategies for combating the COVID-19 pandemic to the institutional and cultural context in their respective countries. In addition to system change, culture change, that is, the establishment of new norms and behavior, is needed.^28^ This change needs to be driven by leaders showing unequivocal and explicit support for outbreak control policies and their implementation, hopefully bringing the outbreak under control and reducing its overall magnitude. This is especially important because the unpredictable future of the pandemic will be exacerbated by public’s misunderstanding of health messages,^29^ causing not only worry but likely also mental health issues in the population.

## Data Availability

The data used for the regression model in this study is available in its entirety in Table 1. The original data sources are referenced in the section Model and methods.

## Human participant protection

No humans participated in this study. The data used for the regression model in this study is available in its entirety in Table 1. The original data sources are referenced in the section Model and methods.

## References

1. Roser M, Ritchi H, Ortiz-Ospina E. Coronavirus disease (COVID-19) - Statistics and research. OurWorldInData.org. https://ourworldindata.org/coronavirus. Published 2020. Accessed March 21, 2020.

2. Hellewell J, Abbott S, Gimma A, et al. Feasibility of controlling COVID-19 outbreaks by isolation of cases and contacts. Lancet Glob Heal. 2020:488–496. doi:10.1016/s2214-109x(20)30074-7

3. Buse K, Dickinson C, Sidibé M. HIV: Know your epidemic, act on its politics. J R Soc Med. 2008;101(12):572–573. doi:10.1258/jrsm.2008.08k036

4. Lowcock EC, Rosella LC, Foisy J, McGeer A, Crowcroft N. The social determinants of health and pandemic H1H1 2009 influenza severity. Am J Public Health. 2012;102(8):51–58. doi:10.2105/AJPH.2012.300814

5. Slusser M. The logic in ecological: I. The logic of analysis. Am J Public Health. 1994;84(5):825–829. doi:doi/10.2105/AJPH.84.5.825

6. Texier G, Farouh M, Pellegrin L, et al. Outbreak definition by change point analysis: A tool for public health decision? BMC Med Inform Decis Mak. 2016;16(1):1–12. doi:10.1186/s12911-016-0271-x

7. Mitjà O, Clotet B. Use of antiviral drugs to reduce COVID-19 transmission. Lancet Glob Heal. 2020;(20):1–2. doi:10.1016/S2214-109X(20)30114-5

8. Institutional Profiles Database (IPD). DG Trésor. http://www.cepii.fr/institutions/EN/ipd.asp. Published 2016. Accessed March 19, 2020.

9. Madhav N, Oppenheim B, Gallivan M, Mulembakani P, Rubin E, Wolfe N. Pandemics: Risks, impacts, and mitigation. In: Jamison DT, Gelband H, Horton S, et al., eds. Disease Control Priorities: Improving Health and Reducing Poverty (Volume 9). 3rd ed. Washington, DC: World Bank Group; 2017:315–345.

10. Imports of Goods and Services (Current US$). The World Bank. https://data.worldbank.org/indicator/NE.IMP.GNFS.CD. Published 2017. Accessed March 19, 2020.

11. Population Density (People per sq. km of Land Area). The World Bank. https://data.worldbank.org/indicator/EN.POP.DNST. Published 2018. Accessed March 19, 2020.

12. Irvin RA, Stansbury J. Citizen Participation in Decision Making: Is It Worth the effort? Public Adm Rev. 2004;64(1):55–65. doi:10.1111/j.1540-6210.2004.00346.x

13. Beierle TC, Konisky DM. What are we gaining from stakeholder involvement? Observations from environmental planning in the Great Lakes. Environ Plan C Gov Policy. 2001;19(4):515–527. doi:10.1068/c5s

14. Santiso C. IMPROVING FISCAL GOVERNANCE AND CURBING CORRUPTION?: HOW RELEVANT ARE AUTONOMOUS AUDIT AGENCIES? 2006;7(2):97–108.

15. Primary Completion Rate, Total (% of Relevant Age Group). The World Bank. https://data.worldbank.org/indicator/SE.PRM.CMPT.ZS. Published 2018. Accessed March 19, 2020.

16. Sorveglianza Integrata COVID-19 in Italia. Ordinanza n. 640, 21 Mar 2020. Istituto Superiore di Sanità. https://www.epicentro.iss.it/coronavirus/bollettino/Infografica_21marzoITA.pdf. Published 2020. Accessed March 22, 2020.

17. Country Comparison: Median Age. CIA World Factbook. https://www.cia.gov/library/publications/the-world-factbook/fields/rawdata_343.txt. Published 2020. Accessed March 19, 2020.

18. Hofstede G. Culture’s Consequences: Comparing Values, Behaviors, Institutions, and Organizations across Nations. 2nd ed. Thousand Oaks, CA: Sage; 2001.

19. Fincher CL, Thornhill R, Murray DR, Schaller M. Pathogen prevalence predicts human cross-cultural variability in individualism/collectivism. Proc R Soc B Biol Sci. 2008;275(1640):1279–1285. doi:10.1098/rspb.2008.0094

20. Borg MA, Camilleri L, Waisfisz B. Understanding the epidemiology of MRSA in Europe: Do we need to think outside the box? J Hosp Infect. 2012;81(4):251–256. doi:10.1016/j.jhin.2012.05.001

21. Mulki JP, Caemmerer B, Heggde GS. Leadership style, salesperson’s work effort and job performance: The influence of power distance. J Pers Sell Sales Manag. 2015;35(1):3–22. doi:10.1080/08853134.2014.958157

22. Winterich KP, Zhang Y. Accepting Inequality Deters Responsibility: How Power Distance Decreases Charitable Behavior. J Consum Res. 2014;41(2):274–293. doi:10.1086/675927

23. P. Vatcheva K, Lee M. Multicollinearity in Regression Analyses Conducted in Epidemiologic Studies. Epidemiology. 2016;06(2):1–9. doi:10.4172/2161-1165.1000227

24. Frank KA. Impact of a confounding variable on a regression coefficient. Sociol Methods Res. 2000;29(2):147–194. doi:10.1177/0049124100029002001

25. Frank KA, Maroulis SJ, Duong MQ, Kelcey BM. What would it take to change an inference? Using Rubin’s causal model to interpret the robustness of causal inferences. Educ Eval Policy Anal. 2013;35(4):437–460. doi:10.3102/0162373713493129

26. Solomon T, Lewthwaite P, Perera D, Cardosa MJ, McMinn P, Ooi MH. Virology, epidemiology, pathogenesis, and control of enterovirus 71. Lancet Infect Dis. 2010;10(11):778–790. doi:10.1016/S1473-3099(10)70194-8

27. Joshi H, Lenhart S, Albright K, Gipson K. Modeling the effect of information campaigns on the HIV epidemic in Uganda. Math Biosci Eng. 2008;5(4):757–770. doi:10.3934/mbe.2008.5.757

28. Ferguson JK. Preventing healthcare-associated infection: risks, healthcare systems and behaviour. Intern Med J. 2009;39(9):574–581. doi:10.1111/j.1445-5994.2009.02004.x

29. Bao Y, Sun Y, Meng S, Shi J, Lu L. 2019-nCoV epidemic: address mental health care to empower society. Lancet. 2020;395(10224):e37–e38. doi:10.1016/S0140-6736(20)30309-3

